# Environmental etiology of psychotic experiences in Ethiopia: Effects of khat use, trauma load and seasonality

**DOI:** 10.64898/2026.09.27.26364112

**Authors:** Michael Odenwald, Laura Oetzel, Hugh F.G.A. Atkinson, Marina Widmann, Zeleke Mekonnen, Matiwos Soboka, Veronika Müller-Bamouh, Sultan Suleman, Fasil Tessema, Stefan W. Toennes, Manuel Mattheisen, Thomas G. Schulze, Markos Tesfaye, Kristina Adorjan

**Author notes:** These authors contributed equally to this work and share first authorship (Michael Odenwald, Laura Oetzel). Corresponding author: Michael Odenwald, Department of Psychology, University of Konstanz, Universitätsstr.10, 78464 Konstanz, Germany.

## Abstract

In Ethiopia, khat (*Catha edulis*) leaves, containing amphetamine-like alkaloids, are chewed for their psycho-stimulating effects. Khat use has been associated with psychosis. In a psychosis etiology framework, we investigated whether season, fluctuations of khat availability and khat use are associated with khat-induced psychotic experiences and whether trauma load moderates this association.

In Southwestern Ethiopia, a cohort of 685 men aged 18-40 years were interviewed twice, during dry and after rainy season (low vs. high khat availability). Khat use was assessed by urine immunoassay and self-report. Trauma load and psychotic experiences were assessed by trained local interviewers.

Khat use increased after the rainy season (*p* < 0.0001). Khat use and seasonality predicted khat-induced psychotic experiences. Main findings comprise increased odds of psychotic experiences among immunoassay-positive individuals with high trauma load. A potential additional interaction with recent trauma emerged. Findings support sensitization-related mechanisms but require replication in larger samples with longer follow-up.

## Introduction

Psychotic disorders are among the most serious mental illnesses. They often place a high burden on patients, their families, and society in general, especially in low- and middle-income countries (LMIC), where few resources are available for treating mental disorders.^1^

While severe psychotic disorders such as schizophrenia represent the extreme end of the psychosis spectrum, subclinical psychotic experiences are considered closer to the healthy state.^2^ Psychotic disorders are relatively rare in the general population, with a lifetime prevalence of approximately 3%.^3^ However, it is important to note that most prevalence data come from high-income countries (HIC).^4^ In comparison, subclinical psychotic experiences are frequent, with a lifetime prevalence of approximately 28%.^5^ An authoritative review revealed that subclinical psychotic experiences are in 75-90% of the cases transient.^6^ Several studies support the continuum model by showing that individuals with subclinical psychotic experiences have a higher risk to experience significant persisting problems in social functioning and develop psychotic disorders later in life.^2^ Environmental factors, such as prenatal stress, malnutrition, infections, trauma, urban upbringing, minority status, and social isolation, are considered important in the etiopathogenesis of psychosis.^7^ Those related to stress or involving dopamine-activating mechanisms may play a key role in its development.^8,9^ Several studies showed that substance-induced psychosis is associated with an elevated risk of later developing schizophrenia (e.g. 25%).^10^

An environmental risk factor that is relatively unique to countries at the Horn of Africa is the chewing of khat (*Catha edulis*) leaves, which is Ethiopia’s most common psychoactive substance.^11^ Problematic khat use is highly prevalent among people with mental disorders.^12^ Excessive use of khat leaves can lead to dependence and khat-induced psychotic experiences or disorders.^11,13^ Khat leaves contain alkaloids, such as cathinone, cathine, and norephedrine (NE), which have stimulating effects on the central nervous system; cathinone (S(-)alpha-aminopropiophenone) is the main psychoactive component with amphetamine-like actions.^14,15^ Cathinone-induced psychological effects are mediated through the mesolimbic dopaminergic pathway; cathinone acts via various mechanisms in the CNS to increase dopamine release.^14^ Given these properties, khat was discussed as a risk factor for the development of psychosis.^16,17^ One unique but poorly studied aspect of khat use as a potential risk factor is its variation in availability and quality across seasons, with higher market availability and pharmacological potency in certain periods.^18^ Whether seasonal shifts also alter consumption levels, potentially via economic dynamics such as price and availability, remains an underexplored gap. Several studies have provided preliminary evidence that the relationship between khat use and psychotic experiences is moderated by posttraumatic stress disorder (PTSD) and trauma load (i.e. number of previously experienced trauma event types).^19^ Odenwald and colleagues^19^ reported in a sample of 8,124 individuals from Somalia that the percentage of respondents with psychotic experiences increased with the amount of khat use and this association was stronger among respondents with than without PTSD.

The present study aimed to add empirical quantitative data on the yet unstudied and poorly understood khat use patterns and their effects at the Horn of Africa. The project aimed to examine, in a representative sample from Ethiopia, the seasonal variation of khat use and the effects of the environmental factors *khat use* and *trauma load* on the development of sub-clinical khat-induced psychotic experiences, a potential vulnerability marker for psychotic disorders.

The present research was guided by three central goals. We sought to determine whether khat use varies between the dry and rainy seasons, two periods in the course of the year with different khat availability (RQ1). Furthermore, we investigated whether the amount of khat use and seasonal fluctuations beyond khat exposure influence the prevalence of sub-clinical khat-induced psychotic experiences (RQ2). Third, we examined whether trauma load moderates the association between khat use and khat-induced psychotic experiences (RQ3). As these research questions have not yet been addressed, we combine a hypothesis-driven with a hypothesis-generating approach.

## Methods

### Study design

The study employs a prospective cohort design. Measurements took place at two timepoints: T1 was in the dry season (February and March 2015), when the supply of khat was limited and khat was expensive. T2 was shortly after the rainy season (November and December 2015), when khat was abundantly available at local markets and prices were low. Trained local interviewers assessed the participants twice (at T1 and T2) to determine the presence and stability of distinct khat-induced psychotic experiences, to quantify khat use and potentially traumatic experiences, and to collect other information on physical and mental health. At both time points, urine samples were collected and assessed for khat alkaloids under field conditions by a simple immunoassay test that had been originally developed to detect amphetamine use. In order to validate our measures, we conducted a validation study involving expert interviews and a more detailed laboratory-based khat alkaloid quantification together with our T1 assessment, which will be reported in detail elsewhere (Preprint: Atkinson and colleagues).^20^

### Study population and recruitment

This study was conducted at the Gilgel Gibe Field Research Center (GGFRC) in Southwestern Ethiopia. The GGFRC is a unique health and demographic surveillance site established and managed by Jimma University (JU). It is located 55 km northeast of Jimma and comprises 8 rural and 3 urban kebeles (the lowest administrative unit in Ethiopia). The center’s registry contains continuously updated data of the total population of roughly 70,000 (30% urban and 70% rural) on sociodemographic, economic, and health variables (https://ju.edu.et/gilgel-gibe-field-research-center/).

We used a two-stage process to randomly select 1,100 male adults from the center’s registry. In the first stage, we randomly chose two urban and three rural clusters. Then, in the second stage, we randomly selected participants based on the overall proportion of the cluster subpopulation size. Inclusion criteria were male sex, age 18-40 years, residency in the GGFRC for at least 6 months and fluency in Amharic or Afaan Oromo. Only men were recruited because in the GGFRC significantly more men than women use khat.^21^ We chose the age range because khat use typically starts in young adulthood, and psychotic experiences usually appear for the first time at a young age.^22^ Exclusion criteria were plans to relocate within the study period, severe alcohol or other substance abuse and the presence of severe developmental and neurological disorders. Since all criteria were already recorded in the registry, random selection was performed only among eligible individuals. The selected participants were then contacted by the local GGFRC enumerators.

The study was approved by the ethics committees of Jimma University and the University of Munich and written informed consent was obtained from all participants. The study was conducted in accordance with the Declaration of Helsinki. This manuscript was prepared in accordance with the Strengthening the Reporting of Observational Studies in Epidemiology (STROBE) guidelines. We briefly reported on this study in a Letter to the Editor.^23^

### Data collection

Local enumerators, experienced GGFRC staff, conducted the interviews. They underwent an initial 5-day training by local and international mental health experts covering the instrument, interviewing rules, role plays, and supervised practice, followed by a 3-day refresher before T2.

### Instruments and measures

#### Self-report of khat use

Khat chewing was assessed with an adapted version of the Timeline Follow back (TLFB) method for the previous week.^24,25^ We created a binary (khat use last week yes vs. no) and a quantitative score, i.e. sum of khat chewing hours in the last week. The binary khat use variable, assessed by trained local interviewers, showed high agreement with expert assessments in the validation study (κ = 0·798).

### Psychotic experiences

We used the information on khat-induced psychotic symptoms from previous population-based studies in khat users^26^ to selected four items of the WHO’s Composite International Diagnostic Interview (CIDI Core Version 2.1, 12 month version): G2, believing that you are being followed; G2b, thinking that people are talking about or laughing at you; G18: hearing things other people could not hear; and G21: having unusual feelings on your skin or inside your body. Based on previous studies, we defined khat-induced psychotic experiences as being present only during or up to 6 hours after the last khat consumption.^26^ Given the transient nature of substance-induced psychotic experiences, we classified experiences as present regardless of whether the respondent recognized their psychotic nature or whether they met the clinical criteria for CIDI coding (e.g., the interviewer’s judgment that the respondent’s explanations were implausible). We calculated a dichotomous variable, *khat-induced psychotic experience,* coded 1 if any of the four items were endorsed and 0 if none. Agreement between local interviewers and experts was moderate (κ = 0·55), with high specificity (0·93) and moderate sensitivity (0·58). Given the moderate agreement, suggesting that most cases and non-cases were correctly classified, we considered the measure adequate for our analysis.

### Trauma Load

Potentially traumatic event types were assessed with the Life Events Checklist for DSM-5 (LEC-5).^24,27^ We assessed only the event types that were either personally experienced by the respondents or directly witnessed. For each of the 17 items, participants responded in a yes/no format to the statement “Happened to me personally, or I witnessed it happen to someone else.” Participants were interviewed with the LEC-5 at T1 and T2. At T1, we asked about trauma event types that had previously happened (*trauma load before T1*), and at T2, we only asked about events from the past nine months since T1 (*recent trauma load*). We defined *lifetime trauma load* as the total sum of event types reported at both time points. Based on this total sum, we split participants into three groups: those who reported no events were classified as *no lifetime trauma load; the* remaining participants were divided into two groups based on median split, i.e. those who reported four or more events as *high lifetime trauma load*, those with one to three events as *low lifetime trauma load*. This categorization logic was subsequently applied to *load before T1*. For recent trauma, anyone who reported at least one event between T1 and T2 was assigned to the recent trauma load group. The rationale for this distinction is grounded in theoretical assumptions that the absence of exposure to any traumatic event type reflects a qualitatively different level of vulnerability to adverse mental health outcomes compared with any level of exposure. In addition, the use of a median-based or comparable threshold approach is a well-established statistical strategy for classifying exposure severity, allowing for the differentiation between low and high trauma load and thereby capturing differences in levels of clinical risk.

### Immunoassay tests

To overcome the methodological weakness of self-report data on khat use we additionally assessed objective pharmacological data. While commercially available rapid urine tests for cathine exist and can indicate recent khat use, there are no field-tested, khat-specific immunoassay kits with peer-reviewed validation data^28^. In our study, khat alkaloids were assessed in the urine by amphetamine immunoassay tests (single test, cut-off 300 ng/mL; nal von minden GmbH, Moers, Germany) at T1, T2 and the validation sub-study. Urine samples were collected in a clean wide-mouth standard urine container with a screw cup. Within hours, they were transported to a local health center and stored in a refrigerator, where trained nurses performed the immunoassay tests. For the validation study (see preprint),^20^ urine samples were subsequently transported to the Jimma University laboratory in a standard cool box and stored at −20°C until analysis by high performance liquid chromatography (HPLC, Agilent 1260 Infinity Serious) at JuLaDQ.^28^ NE was selected as the single reference substance for practical reasons (longer detection window and legal availability of the pure substance in Ethiopia). NE intake through cold medications was controlled for. Agreement between measures (n = 119) was moderate (κ = 0·43). Specificity was perfect (1·00), but sensitivity was low (0·52), indicating limited detection of low-level khat use. Thus, the immunoassay is most suitable for screening heavy, recent khat use. Participants with positive immunoassay tests at T1 reported increased chewing hours in the last week than those with negative results (negative: *M* = 5·25, *SD* = 8·55 vs. positive: *M* = 15·49, *SD* = 15·35, *t*(232·22) = −8·61, *p* < 0·0001, *Hedges’ g* = −0·94).

### Statistical analysis

We used R (https://www.r-project.org/) to evaluate group differences, applying McNemar’s test for binary khat-use outcome variables and two-sided paired-samples *t*-tests for the continuous khat-use outcome variable. Because the normality assumption was not met, as assessed by the Shapiro-Wilk test and visual inspection of Q-Q plots, the nonparametric Wilcoxon signed-rank test with continuity correction was additionally performed to confirm the robustness of the results. Effect sizes were calculated using Cohen’s *d* for paired-samples comparison (standardized mean difference based on paired observations) and Cohen’s *g* for McNemar’s tests, calculated as *g* = *b*/(*b* + *c*) − 0.5, where *b* and *c* represent the two types of discordant pairs. The Bonferroni correction was applied to control for alpha error accumulation.

Mixed-effects binary logistic regression models were estimated using the lme4 package to examine the associations of seasonality and khat use with khat-induced psychotic experiences. Random intercepts were specified to account for repeated observations of the dependent and independent variables at T1 and T2. In addition, separate binary logistic regression models, using the rms-package in R, were conducted for T1 and T2 using the same predictors and outcome. These timepoint-specific models were run for all participants with available data at that timepoint (see Supplementary Material).

To analyze lifetime trauma load as a time-invariant moderator (RQ3; reference category: high lifetime trauma load), a mixed-effects logistic regression model with random intercepts for repeated measurements and separate binary logistic regression models for T1 and T2 were estimated. For the dry-season model, ordinal trauma load assessed before T1 was included, whereas the rainy-season and mixed-effects models incorporated the ordinal lifetime trauma load variable. Based on prior validity testing of the immunoassay and the results from RQ1, moderation analyses included binary khat use predictors, as binary and continuous operationalizations yielded comparable patterns of results. We chose the biological khat predictor, as this provides an objective measure of recent high use. Chewing hours were not further investigated as a predictor, as it represents an imprecise proxy for actual exposure and does not adequately reflect variability in dose. The highest trauma category was specified as the reference level across models to facilitate interpretation of immunoassay effects under conditions of greatest vulnerability. To evaluate the overall contribution of predictors and their interaction, Type III Wald χ^2^ tests were computed for fixed effects. This approach tests each term in the model while adjusting for all other included predictors. Significant interaction effects were further examined using estimated marginal means (EMMs), allowing comparison of immunoassay effects across levels of lifetime trauma load. Full model results are reported in the Supplementary Material.

Finally, to investigate a potential three-way interaction between immunoassay test results at T2 (binary: yes vs. no), trauma load before T1 (binary: low vs. high), and recent trauma load (binary: yes vs. no) on T2 khat-induced psychotic experience, a binary logistic regression was conducted. To reduce instability in our trauma predictor variable due to low prevalence, participants with no trauma load before T1 were excluded from the analysis.

The significance level (α) was set at 0·05 for all statistical analyses.

Assumption testing was conducted for all analyses. Model assumptions were evaluated prior to analysis, and where violations were identified, appropriate alternative statistical methods were applied, as described above. All other assumptions were met.

Given the low proportion of missing data (≤4%), except for hours of khat chewing in the past week, and the robustness of mixed-effects logistic regression to missingness,^29^ missing values were handled within the models or by listwise deletion. For hours of khat chewing, Little’s MCAR test was non-significant (χ^2^(20) = 18·60, p = 0·55), supporting the use of listwise deletion.

### Code availability

The underlying analytical code for this study is not publicly available but may be made available to researchers upon reasonable request from the corresponding author.

## Results

Of the randomly selected 1,100 individuals, 853 (77·5%) agreed to participate at T1. A total of 685 participants (81·5%) completed the T2 assessment; our analyses will be based on this completer sample. For details on participant flow and reasons for exclusion, see Figure S1 and Supplementary Eligibility Criteria. Their average age was 28.6 years (SD = 6·6, median = 29), with 213 residing in urban and 472 in rural areas.

A detailed overview of the dropout analysis is provided in Table S1. Only a younger age was significantly associated with drop-out at T2 (*p* = 0·015).

### Descriptive statistics

Khat use was very common among participants at T1 and T2 with around a third chewing considerable amounts causing a positive immunoassay and one in ten reporting khat-induced psychotic experiences (see Table 1).

**Table 1.**
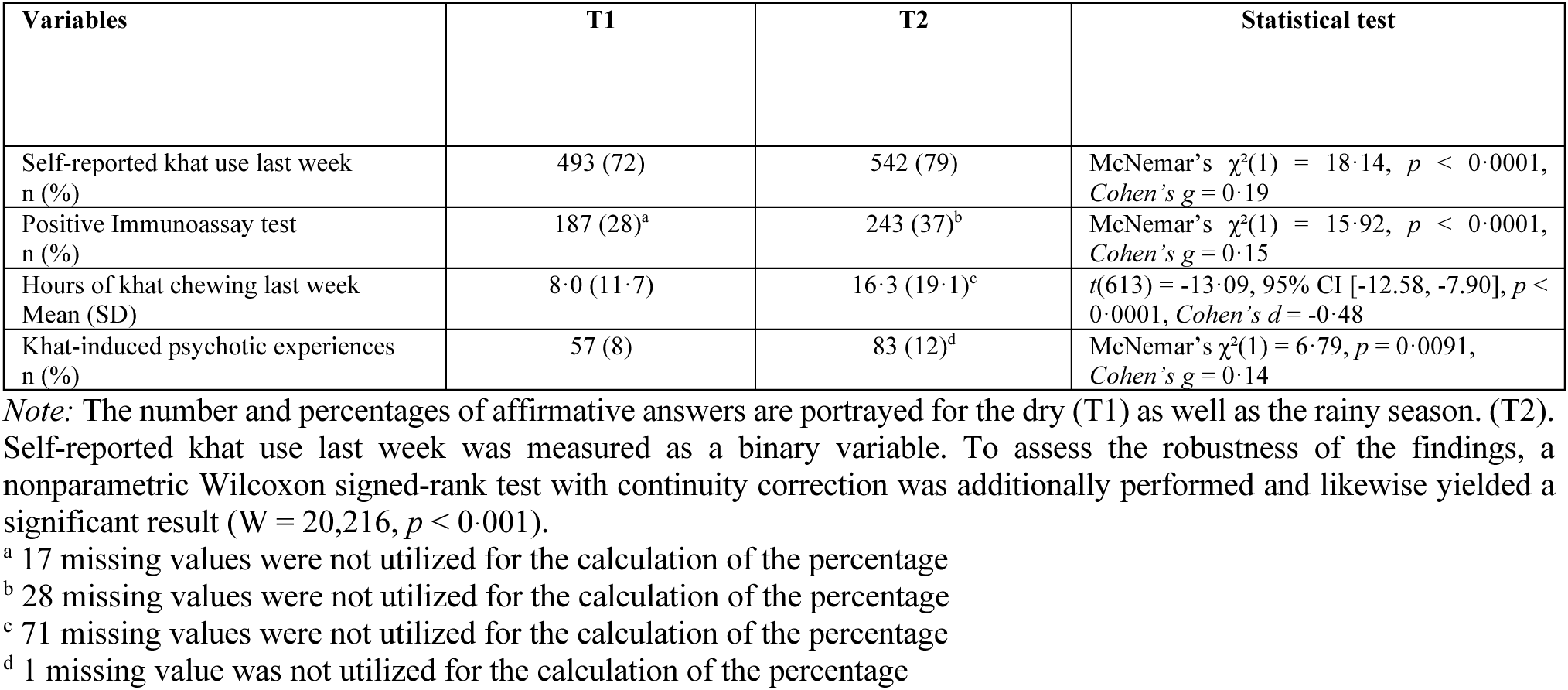
Descriptives of the participants who completed both assessments (N = 685)

| Variables | T1 | T2 | Statistical test |
| --- | --- | --- | --- |
| Self-reported khat use last week<br>n (%) | 493 (72) | 542 (79) | McNemar's $\chi^2(1) = 18.14, p < 0.0001$,<br>*Cohen's g* = 0.19 |
| Positive Immunoassay test<br>n (%) | 187 (28)<sup>a</sup> | 243 (37)<sup>b</sup> | McNemar's $\chi^2(1) = 15.92, p < 0.0001$,<br>*Cohen's g* = 0.15 |
| Hours of khat chewing last week<br>Mean (SD) | 8.0 (11.7) | 16.3 (19.1)<sup>c</sup> | $t(613) = -13.09, 95\% \text{ CI } [-12.58, -7.90], p < 0.0001, \text{Cohen's } d = -0.48$ |
| Khat-induced psychotic experiences<br>n (%) | 57 (8) | 83 (12)<sup>d</sup> | McNemar's $\chi^2(1) = 6.79, p = 0.0091$,<br>*Cohen's g* = 0.14 |

*Note:* The number and percentages of affirmative answers are portrayed for the dry (T1) as well as the rainy season. (T2). Self-reported khat use last week was measured as a binary variable. To assess the robustness of the findings, a nonparametric Wilcoxon signed-rank test with continuity correction was additionally performed and likewise yielded a significant result ( $W = 20,216, p < 0.001$ ).
<sup>a</sup> 17 missing values were not utilized for the calculation of the percentage
<sup>b</sup> 28 missing values were not utilized for the calculation of the percentage
<sup>c</sup> 71 missing values were not utilized for the calculation of the percentage
<sup>d</sup> 1 missing value was not utilized for the calculation of the percentage

A total of 74 men reported no lifetime trauma load, while 287 reported low and 323 high trauma load (one participant due to a missing T2 LEC assessment). The average number of lifetime trauma event types was 3·7 (SD = 2·7, median = 3).

323 participants experienced low trauma load before T1, while 272 reported high trauma load before T1. The average number of trauma event types before T1 was 3·2 (SD = 2·4, median = 3). Recent trauma load was less common: 204 participants experienced at least one trauma event type between T1 and T2, while 480 reported none.

### Research question 1: Does khat use differ between dry and rainy season?

The rainy season was associated with a significant increase in khat use compared to the dry season (see Table 1), for self-reported use, immunoassay results and self-reported hours of khat chewing. All *p*-values remain significant after Bonferroni correction for the three comparisons (adjusted α = 0·0167).

### Research question 2: Do khat use and season affect the prevalence of khat-induced psychotic experiences?

In the mixed-effects logistic regression model, both season and immunoassay results significantly predicted khat-induced psychotic experiences (season: Est. = 1·07, SE = 0·36, *p* = 0·00299; immunoassay: Est. = 1·44, SE = 0·45, *p* = 0·0012; R^2^ marginal = 0·01, R^2^ conditional = 0·95); results are depicted in Figure 1. Comparable findings emerged when using self-reported khat use in the past week as a predictor (season: Est. = 1·2, SE = 0·35, *p* = 0·00056; self-report: Est. = 1·67, SE = 0·72, *p* = 0·0195; R^2^ marginal = 0·02, R^2^ conditional = 0·95). These results were supported by separate logistic regressions (see Tables S2 - S4).

**Figure 1.**
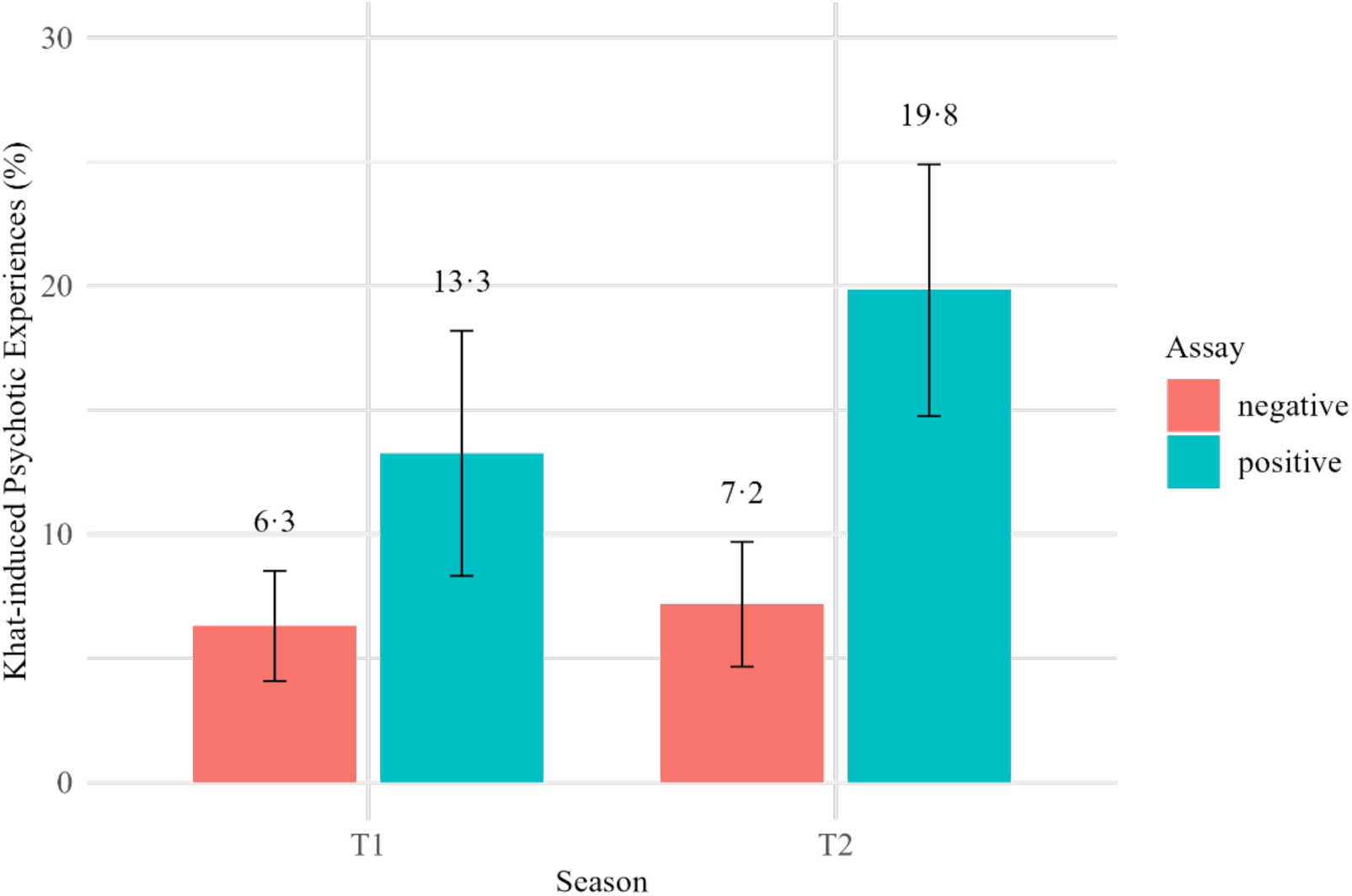
Percentage of participants with khat-induced psychotic experiences by assay and season. ***Note.*** *N =* 641 (T1/Assay negative *n* = 460; T1/Assay positive *n* = 181; T2/Assay negative *n* = 404; T2/Assay positive *n* = 237). The percentages of khat-induced psychotic experiences are shown according to immunoassay results and season. The error bars represent 95% confidence intervals.

### Research question 3: Do trauma load and khat use interact to predict khat-induced psychotic experiences?

Even though lifetime trauma load (time-invariant) was not a significant predictor, its interaction with immunoassay results was significant in the mixed-effects model (Est. = −3·55, *SE* = 1·26, *z* = −2·82, *p* = 0·0048, R^2^ marginal = 0·02, R^2^ conditional = 0·96). Detailed results, including separate logistic models for T1 and T2, are provided in the Supplementary Material (Table S5, S8 − S9).

Type III Wald χ^2^ tests confirmed a significant interaction between immunoassay and lifetime trauma load (χ^2^ = 7·95, p = 0·019), with no main effect of trauma observed (see Table S6). To interpret this interaction, estimated marginal means were used to examine immunoassay effects within each trauma level. These showed a significant association only in the high-trauma group (Est. = −2·68, SE = 0·69, z-ratio = −3·91, p = 0·0001), with no significant differences in the low- or medium-trauma groups (see Table S7). In the high-trauma group, immunoassay positivity was associated with higher odds of khat-induced psychotic experiences. Figure 2 shows a graphical display of immunoassay status and trauma load before T1 (dry season) and lifetime trauma load (rainy season).

**Figure 2.**
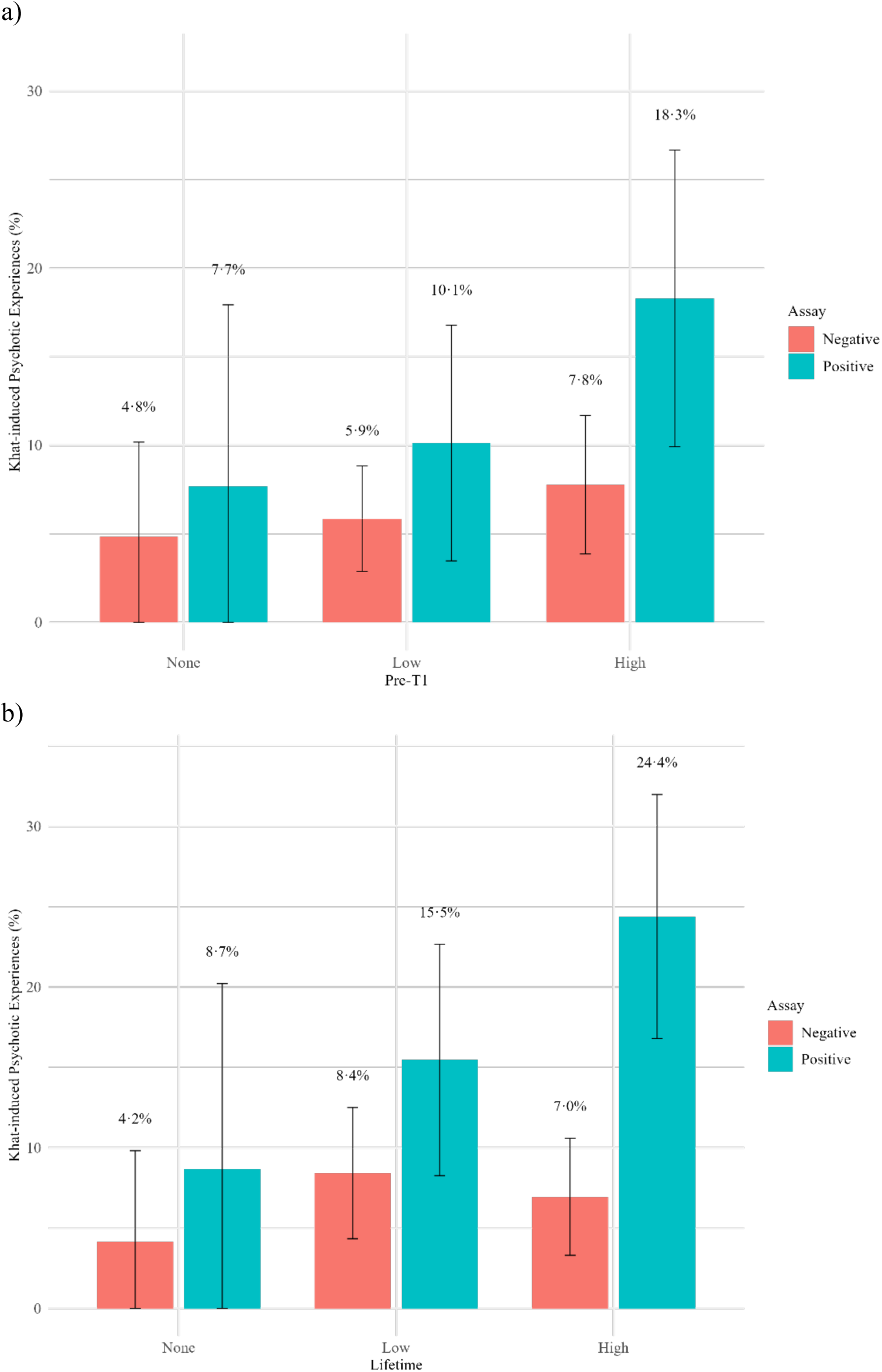
Percentage of participants with khat-induced psychotic experiences by assay and trauma load. a) T1 model (dry season) using trauma load before T1 b) T2 model (rainy season) using lifetime trauma load ***Note.*** *N =* 668 (dry)/665 (rainy) (Pre-T1 None/Assay Negative *n* = 62; Pre-T1 None/Assay Positive *n* = 26; Pre-T1 Low/Assay Negative *n* = 239; Pre-T1 Low/Assay Positive *n* = 79; Pre-T1 High/Assay Negative *n* = 180; Pre-T1 High/Assay Positive *n* =82; Lifetime None/Assay Negative *n* = 48; Lifetime None/Assay Positive *n* = 23; Lifetime Low/Assay Negative *n* = 178; Lifetime Low/Assay Positive *n* = 97; Lifetime High/Assay Negative *n* = 187; Lifetime High/Assay Positive *n* = 123). The percentages of khat-induced psychotic experiences at T1 and T2 are shown according to immunoassay results and trauma load before T1/ lifetime trauma load. The error bars represent 95% confidence intervals.

The three-way interaction between the independent variables immunoassay result, trauma load before T1 and recent trauma load did not reach significance (*b* = −2·0, *SE* = 1·13, *z* = −1·76, *p* = 0·079, McFadden’s pseudo-R^2^ = 0·06). More details on this model can be found in Table S10.

Even though the logistic regression models did not reveal a statistically significant interaction, the visual inspection of the bar chart in Figure 3 suggests a potential interaction effect. When trauma load before T1 is low, recent load seems to be associated with an increased percentage of khat-induced psychotic experiences among high khat users, resulting in a similar percentage of khat-induced psychotic experiences as in groups with high trauma load before T1.

**Figure 3:**
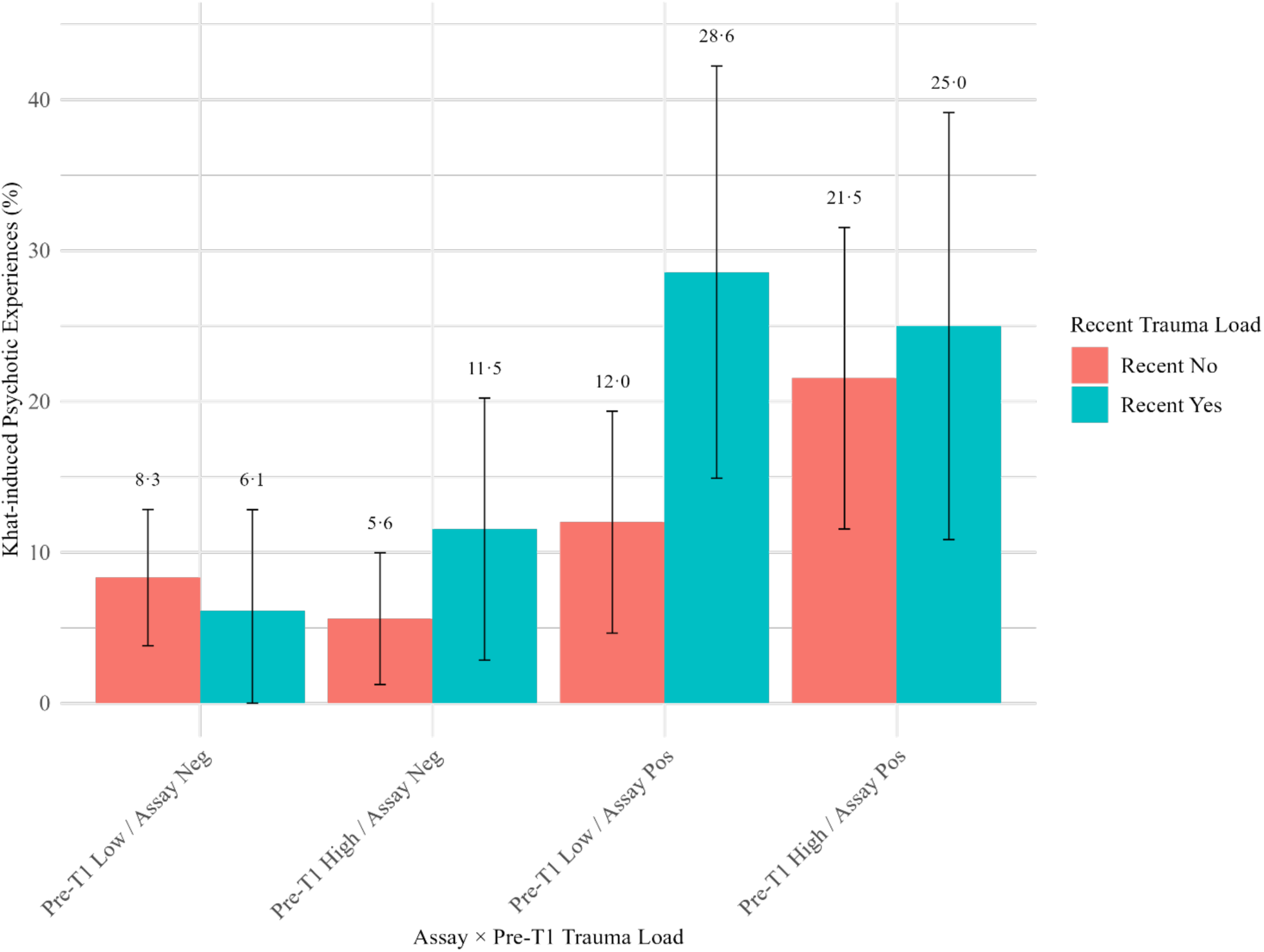
Percentage of participants with khat-induced psychotic experiences by assay, trauma load before T1 and recent trauma (restricted to participants with trauma load before T1) *Note. N =* 573 (Pre-T1 Low/Assay Neg/Recent No *n* = 144; Pre-T1 Low/Assay Neg/Recent Yes *n* = 49; Pre-T1 High/Assay Neg/Recent No *n* = 107; Pre-T1 High/Assay Neg/Recent Yes *n* = 52; Pre-T1 Low/Assay Pos/Recent No *n* = 75; Pre-T1 Low/Assay Pos/Recent Yes *n* = 42; Pre-T1 High/Assay Pos/Recent No *n* = 65; Pre-T1 High/Assay Pos/Recent Yes *n* = 36). The percentages of khat-induced psychotic experiences at T2 are shown according to immunoassay results, trauma load before T1 and recent trauma load. The error bars represent 95% confidence intervals.

## Discussion

This study shows that khat use varies between times of the year with different khat availability. Furthermore, our data provide preliminary evidence that khat-induced psychotic experiences may be influenced by three environmental factors: 1) individual khat use, 2) seasonal variations beyond khat availability and 3) trauma load. Both objective and self-report measures indicated higher khat consumption and longer chewing duration after the rainy season, when khat availability increases, compared to the dry season. Moreover, after the rainy season, we found a higher prevalence of khat-induced psychotic experiences. Additional analyses provided evidence for complex interaction effects between khat use and the timing of trauma load.

Our study confirms for the first time with quantitative data that the consumption of khat is strongly influenced by its seasonal availability, suggesting that the season during which assessments are conducted should be considered in future research. Moreover, the findings give rise to the theoretical assumption that environmental risk factors for mental disorders may not be static but oscillate across different seasons, in the case of our study site with a higher prevalence of experiencing psychotic phenomena in the weeks following the rainy season an increased risk for developing psychotic symptoms. Beyond the higher khat availability, at our project site there are marked seasonal fluctuations of food security, environmental temperature or labor work demand, factors that can be termed as environmental stressors. Furthermore, a large unwritten knowledge on potency of khat, related to different types and seasonality is locally existing that has not been targeted by scientific research.^18^ In this context, future research should aim to disentangle the potential mental health effects attributable to changes in substance availability and potency, while also considering other social and environmental factors associated with seasonality. Both objective and self-reported khat use were linked to psychotic experiences, reinforcing evidence of its adverse effects in a large male sample.

A significant interaction was observed between analytically detectable khat alkaloids in urine and lifetime trauma load. Estimated marginal means indicated that the association between immunoassay positivity and psychotic experiences was restricted to the high-trauma group, with no effects in lower trauma groups. This pattern suggests that elevated lifetime trauma load may increase vulnerability to khat-induced psychotic experiences following recent khat use.

Further descriptive analysis, dividing lifetime trauma load into load before T1 and recent load, suggested another interaction effect: Among participants with low trauma load before T1 and a positive immunoassay, recent trauma load was associated with an increased percentage of khat-induced psychotic experiences. Although not statistically significant, this points to a potential role of trauma timing and accumulation, with recent trauma contributing particularly when prior trauma is low. The relevance of trauma timing for psychotic symptoms has been shown,^30^ but its interaction with substance use and khat-induced psychotic phenomena remains understudied.

This is one of the first studies to explore this stimulant × trauma interaction in a representative sample of a LMIC, with potential relevance for countries where stimulant consumption and trauma exposure are widespread (e.g. Golden Triangle, South Africa). The findings align with prior research showing stronger associations between khat use and psychotic outcomes in individuals with high trauma load or PTSD versus those with low trauma load.^19^ Also, in HIC, stimulant × stress interaction has been associated with an increased risk of psychotic phenomena,^31^ but evidence is inconclusive, partly due to the small number of users, underscoring the value of studying these associations in traditional khat-consuming populations.

This hypothesized interaction effect aligns with the sensitization model of psychosis that has rarely been studied in humans.^8^ Additional support comes from a broader sensitization approach that studies the interaction of environmental risk factors^32^ and longitudinal and cross-sectional studies showing that childhood trauma exposure combined with later tetrahydrocannabinol (THC) use, increases the risk of psychotic experiences or psychotic disorders.^33^

Our study potentially adds new aspects to the validity of the sensitization hypothesis: First, we found that in the general population, not just high-risk subgroups, the interaction of the environmental risk factors *stimulant use* and *trauma load* may influence the development of khat-induced psychotic experiences, similar to the interaction between cannabis and trauma.^33^ Second, there may be cross-sensitization of prior trauma exposure and stimulant use (again like cannabis). In the Horn of Africa, where millions of people use khat daily, the question of whether a sensitization-like interaction exists between two common environmental risk factors carries a significant public health relevance. Future studies with adequate methods and power are needed to address the current lack of evidence. Research should examine whether khat-induced psychotic experiences reflect underlying mental health conditions or result from sensitization processes, and clarify the role of trauma, particularly adverse childhood experiences and age at exposure, in the development of psychotic experiences.^30^

Furthermore, this study highlights the need for more research on khat use and its psychiatric consequences, as potential severe mental health harms may be underrecognized due to limited data from khat-consuming countries, which is reflected in current evaluation of substance-related risks.^34^ High exposure to environmental stressors, such as food insecurity, violence and lack of medical care, and the high prevalence of psychosis in disadvantaged populations call for adequate prevention and treatment tools in khat countries.^12,35^

Although these findings are exploratory and require further validation, it justified to discuss possible practical consequences, given the public health relevance. Clinics should systematically monitor seasonal variations in demand for mental health services, and programs should target multiple, interacting risk factors by combining trauma-informed care, substance use prevention, and early detection of psychotic symptoms. The complex interplay of these factors highlights the need for complementary care frameworks that add new aspects to established approaches such as the World Health Organization’s mhGAP.^1^ Based on our preliminary analyses, mental health programs in these regions may benefit from seasonally informed approaches, including increased screening, psychosocial support, diverse interventions and community education during periods of higher khat use, such as after the rainy season. Such efforts could potentially be strengthened by training local community health workers to identify high-risk individuals, as illustrated in this study, and by developing culturally and regionally adapted tools that account for environmental stressors.

These findings also highlight the potential to advance our understanding of psychosis onset through gene × environment interaction research.^7^ Populations with widespread stimulant use and high levels of psychosocial stress offer a unique context for identifying specific risk factors and exploring these interactions. Future studies should apply genetically informed, population-based epidemiological designs to investigate gene × environment interplay, in order to develop more targeted, context-specific prevention and intervention strategies and deepen our understanding of basic mechanisms of psychosis development. Longitudinal studies incorporating clinical outcomes will be essential to clarify these pathways and evaluate their public health impact.

One major strength of this study lies in its longitudinal design, which enabled the investigation of temporal associations while accounting for seasonal variation in khat consumption and availability. This design provided insights into how environmental context may shape substance use and mental health outcomes and generated hypotheses for future research.

In addition, this study is among the first to apply this design to investigate khat use in relation to trauma history, offering new insights into how these factors may interact to influence psychotic experiences.

The inclusion of a representative sample from an understudied population further enhances the generalizability of the findings and addresses a significant gap in the literature on substance use and mental health in low-income, high-adversity settings. The general population of countries around the Horn of Africa is highly exposed to environmental risk factors, e.g. a high prevalence of exposure to multiple trauma event types and stimulant use^36^.

Another strength of the study lies in its use of measurement methods well adapted to field conditions. We applied a methodology that partly accounts for the complexity of khat use, i.e. a quantification of recent use and a mix of self-report and objective measures. Compared to ratings by mental health experts, the assessment of khat use by trained local interviewers showed high validity and their assessment of khat-induced psychotic experiences was acceptable. Taken together, these findings support the reliability and feasibility of employing trained local interviewers to assess both environmental risk factors and psychotic experiences in community-based field settings; nevertheless, more resources would have been needed to further improve screening capacities for psychotic phenomena.

The study also had some limitations. First, we did not assess clinical disorders, e.g. we did not directly screen for PTSD and only measured the number of trauma event types experienced by participants. We also did not evaluate psychotic disorders as an outcome variable. Additionally, data on comorbid disorders, other forms of substance use or broader mental health issues were not collected. This needs to be remedied in future studies with a larger sample size and a longer observation period that include such measures.

The biological screening methods in this study showed only moderate-to-low sensitivity. However, validation against highly specific reference standards allowed hypothesis testing. The immunoassay primarily detected high or recent khat use,^28^ especially after the rainy season when participants chewed more hours per week. Many users still fell below the positive threshold, indicating the test mainly identifies high or excessive use. Future research should develop and validate more sensitive tools for detecting low to moderate khat use in field conditions.

Another limitation is that study dropouts were not entirely random; those who left were significantly younger than participants who completed both assessments.

Our study included only male participants, as khat use is predominantly male and female use remains highly stigmatized in rural areas such as our setting. Future studies should include sex to identify general mechanisms, though recruiting women will require targeted strategies to overcome barriers documented in prior research.^24^ We did not distinguish between within-person and between-person effects of khat use. Future research should disentangle these components to clarify whether associations with khat-induced psychotic experiences reflect changes within individuals over time or stable differences between individuals.

Finally, the study may have been underpowered to detect some effects. In particular, the relatively small number of participants with no lifetime trauma load likely reduced precision of trauma-related effect estimates, as reflected in larger standard errors.^29,37^ This was also evident in the three-way interaction between no trauma load before T1, immunoassay results and recent load, which involved the smallest subgroups. Additionally, among the remaining three-way interaction groups (excluding the no trauma load before T1 group), sizes varied up to 3.5-fold, likely reducing precision of effect estimates, reflected in larger standard errors. This issue is especially important given the relatively low prevalence of khat-induced psychotic experiences.

In the absence of a standardization of measuring khat use comparable to the “standard drink” definition of alcohol, studies on khat and its effects can’t avoid using a simplistic approach to overcome the complexity caused by varieties and the absence of stable qualities across seasons.

## Data availability

De-identified individual participant data, including a partial data dictionary, underlying the results reported in this article (including text, tables, figures and appendices) will be made available under certain conditions. Participant consent for data sharing was not explicitly obtained at the time of study. Therefore, access is subject to restrictions.

Data will be released following the publication of a related manuscript currently in preparation, with no predefined end date for availability thereafter.

Access to the data will be granted on a case-by-case basis upon reasonable request by investigators who provide a methodologically sound research proposal, which will be reviewed by a review committee. Data sharing is intended for the purpose of individual participant data meta-analyses.

Approved requests will require a signed data access agreement. Data will then be provided via a secure private link, valid for up to two years.

All proposals should be directed to.

## Acknowledgements

This study was the first to be performed as a joint project by Jimma University located in Jimma, Ethiopia, the Institute of Psychiatric Phenomics and Genomics (IPPG), University Hospital, LMU, in Munich, Germany, and the University of Konstanz in Konstanz, Germany. We thank all study participants and members of the local research team. Furthermore, we want to thank Mustafa al’Absi (University of Minnesota, USA) for his valuable input, which significantly contributed to the interpretation of our data. We also thank Heike Riedke (Konstanz, Germany) for her support in the training of Ethiopian staff on immunoassay testing, sample handling and interpretation.

The project was funded by the Lisa Oehler Foundation and the University of Konstanz (Committee on Research, AFF). The funders had no role in study design, data collection and analysis, decision to publish or preparation of the manuscript.

## Author contributions

MO led the conceptualization, investigation and was involved in data curation, methodology, formal analysis and visualization, while also securing funding and overseeing project administration, resources, supervision and validation. LO was responsible for data curation, methodology, formal analysis and visualization. HA took part in data curation, methodology and formal analysis. MW carried out the investigation and supported project administration. ZM was involved in investigation and methodology and contributed resources. MS took part in the investigation and contributed to methodology and project administration. VMB was involved in the investigation. SS participated in investigation and methodology and provided resources. FT contributed to conceptualization and methodology and was responsible for data management. SWT contributed methodological expertise and was involved in supervision and validation. MM was involved in methodology, supervision and validation. TGS contributed to methodology, secured funding and was involved in supervision and validation. MT participated in methodology, supervision and validation. KA led the conceptualization and contributed extensively to data curation, methodology, formal analysis and visualization, while also securing funding and overseeing project administration, resources, supervision and validation.

MO, LO, HA and KA took the lead in drafting the manuscript and integrating the co-authors’ feedback. The remaining co-authors critically reviewed the manuscript and contributed important intellectual content.

All authors confirm that they had full access to all study data, take responsibility for the decision to submit the manuscript for publication and approve the final version for publication.

