## Supplemental Material for "Environmental etiology of psychotic experiences in Ethiopia: Effects of khat use, trauma load and seasonality"

### Supplementary Material

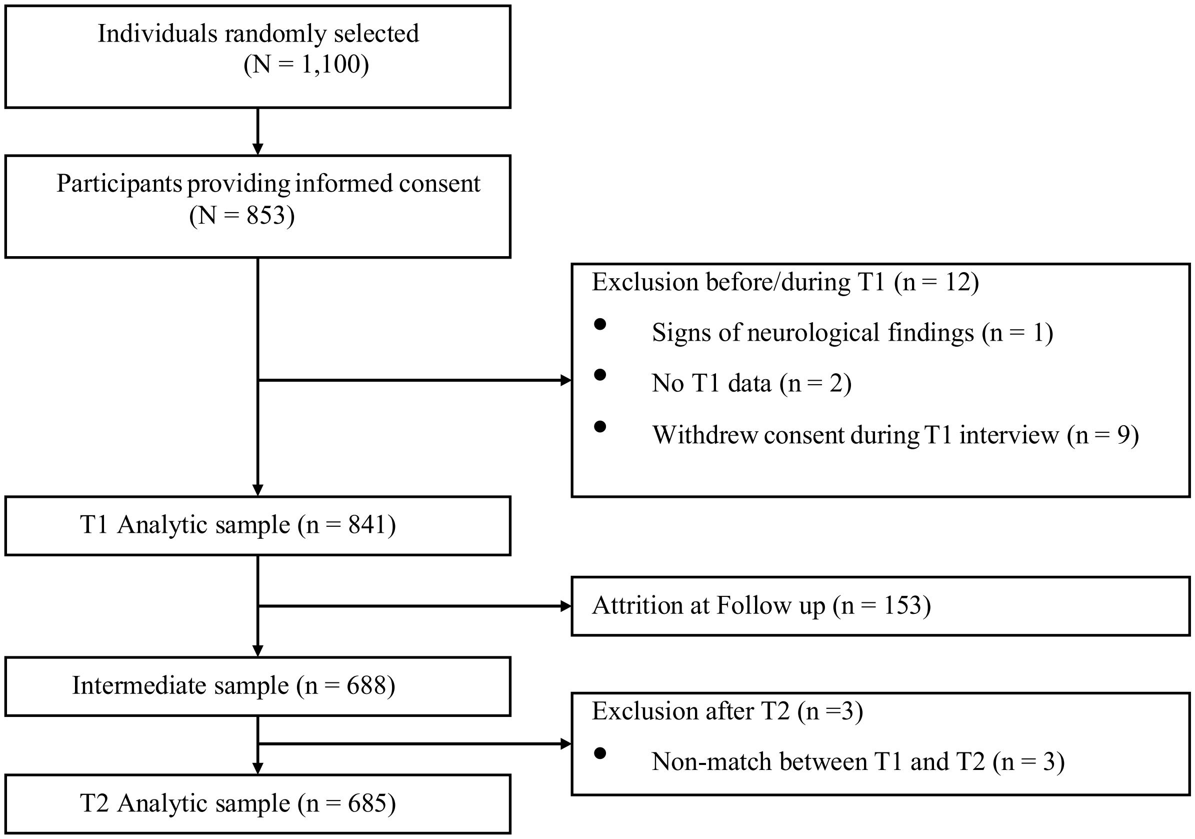

**Figure S1: Participant flow and exclusions in the khat validation study**
*Note.* The flowchart illustrates the number of participants progressing through the study stages.

**Supplementary Eligibility Criteria: Rules for exclusion and inclusion after completed T1 and T2 assessments**

After the participant matching for T1 and T2 was completed, a re-evaluation was conducted. The aim was to verify the accuracy of the matching process based on the individual IDs at T1 and T2. Two types of mismatches were identified: (1) a T1 ID could not be found in the T2 dataset and the matched T2 ID appeared twice, indicating that the ID was correctly matched to one T1 ID but incorrectly assigned to another (double entry); and (2) a T1 ID could not be found in the T2 dataset and the matched T2 ID appeared only once (single entry).

- **Double entry:** If neither location ID nor house number match, then omit this participant, even if names of T1 and T2 might be similar. Note: In these cases, two people in T2 with same location ID, house number and individual ID occurred and severe confounding could not be ruled out.
- **Double entry:** If at least two out of the three variables match and the other variable only shows a slight deviation in T2 from T1, i.e. one number differs, then it is a match.
- **Single entry:** If house number and location ID match and name is the same or similar at both timepoints, then it is a match.
- **Single entry:** If in location ID and/ or house number in T2 compared to T1 only a slight difference occurs, i.e. one number differs or two numbers are switched, and the name of the participant is the same or similar at both timepoints, then it is a match.
- **Single entry:** If location ID does not match, but house number is the same between T1 and T2 as well as the participants name is the same or similar at both timepoints, then it is a match.
- **Single entry:** To minimize data entry errors, two T2 datasets containing the same set of participants were independently entered by two different entry clerks. If, across these two T2 datasets, two records, one from each dataset, were assigned slightly different names and individual IDs but shared matching house numbers, location IDs and sticker IDs, the records were considered to originate from the same individual. Matching to T1 was then performed according to the rules described above.

**Table S1: Dropout analysis: Binary logistic regression model**

| Predictor | *b* | *SE* | *z* | *p* |
|---|---|---|---|---|
| (Intercept) | -0·65 | 0·41 | -1·61 | 0·11 |
| Age | -0·03 | 0·01 | -2·44 | 0·015 |
| Physical_health | -0·01 | 0·05 | -0·10 | 0·92 |
| T1_trauma | 0·04 | 0·04 | 1·19 | 0·23 |
| MH_help | 0·34 | 0·75 | 0·45 | 0·65 |
| MH_function | -0·65 | 0·63 | -1·02 | 0·31 |

*Note.* N = 839. Physical_health = sum of physical complaints; T1_trauma = trauma event types before T1; MH_help = mental health assistance sought; MH_function = mental health related problems in functioning. *b* represents unstandardized regression weights. For logistic regression these are logits, also named log-odds. The model has a null deviance (835) of 792·76 and a residual deviance (830) of 784·70. Due to participant drop-out after T1, this regression was conducted to identify predictors of drop-out with the following variables being included simultaneously in the model: participant age at T1, prior use of mental health services, mental health-related functional impairment, the sum of reported physical complaints and the sum of potentially trauma event types before T1.

**Table S2: Mixed-effects logistic models for RQ2 with predictors a) amphetamine and b) self-reported khat use last week**

a)

| Fixed Effects | | | | |
|---|---|---|---|---|
| | Est | SE | z | p |
| (Intercept) | -8·9 | 0·71 | -12·5 | < 0·0001 |
| season | 1·07 | 0·36 | 2·97 | 0·00299 |
| assay | 1·44 | 0·45 | 3·23 | 0·0012 |
| **Random Effects** | | | | |
| | | | Variance | SD |
| individ (Intercept) | | | 62·84 | 7·93 |
| **Model fit** | | | | |
| $R^2$ | | | Marginal | Conditional |
| | | | 0·01 | 0·95 |

b)

| Fixed Effects | | | | |
|---|---|---|---|---|
| | Est | SE | z | p |
| (Intercept) | -9·63 | 0·95 | -10·17 | < 0·0001 |
| season | 1·2 | 0·35 | 3·45 | 0·00056 |
| Self-report | 1·67 | 0·72 | 2·3 | 0·0195 |
| **Random Effects** | | | | |
| | | | Variance | SD |
| individ (Intercept) | | | 57·54 | 7·59 |
| **Model fit** | | | | |
| $R^2$ | | | Marginal | Conditional |
| | | | 0·02 | 0·95 |

Note. N = 683 (amphetamine model) / 685 (self-report model). season = season variable for T1 and T2; assay = immunoassay result; individ = individual ID; self-report = self-reported khat use last week. Reference categories are negative for the immunoassay test and no use for self-report. p-values for fixed effects calculated using Satterthwaites approximations. Confidence Intervals have been calculated using the Wald method. Model equation: cidi ~ season + assay + (1 | individ) and cidi ~ season + self-report + (1 | individ)

**Table S3: Binary logistic regression models for dry season T1 for RQ2 using predictors a) immunoassay and b) self-reported khat use last week**

a)

| Predictor | *b* | *SE* | *z* | *p* |
|---|---|---|---|---|
| (Intercept) | -2·68 | 0·19 | -14·41 | < 0·0001 |
| T1_assay | 0·81 | 0·28 | 2·84 | 0·0045 |

b)

| Predictor | *b* | *SE* | *z* | *P* |
|---|---|---|---|---|
| (Intercept) | -3·27 | 0·39 | -8·5 | < 0·0001 |
| T1_self | 1·09 | 0·41 | 2·65 | 0·0081 |

Note. N = 668 (amphetamine model)/ 685 (self-report model). T1_assay = immunoassay for T1; T1_self = self-reported khat use in the last week. Reference categories are negative for the immunoassay test and no use for self-report. *b* represents unstandardized regression weights. For logistic regression these are logits, also named log-odds. The amphetamine model has a null deviance (667) of 384·81 and a residual deviance (666) of 377·06. The self-report model has a null deviance (684) of 392·57 and a residual deviance (683) of 383·7. The amphetamine model explained 2·0% of the variance and the self-report model explained 2·26%, based on McFadden's pseudo-$R^2$.

**Table S4: Binary logistic regression models for rainy season T2 for RQ2 using predictors a) immunoassay and b) self-reported khat use last week**

a)

| **Predictor** | ***b*** | ***SE*** | ***z*** | ***p*** |
|---|---|---|---|---|
| (Intercept) | -2·55 | 0·19 | -13·43 | < 0·0001 |
| T2_assay | 1·12 | 0·25 | 4·48 | < 0·0001 |

b)

| **Predictor** | ***b*** | ***SE*** | ***z*** | ***p*** |
|---|---|---|---|---|
| (Intercept) | -3·12 | 0·42 | -7·48 | < 0·0001 |
| T2_self | 1·32 | 0·43 | 3·04 | 0·0024 |

Note. N = 656 (amphetamine model)/ 684 (self-report model). T2_assay = immunoassay for T2; T2_self = self-reported khat use in the last week for T2. Reference categories are negative for the immunoassay test and no use for self-report. *b* represents unstandardized regression weights. For logistic regression these are logits, also named log-odds. The amphetamine model has a null deviance (655) of 474·51 and a residual deviance (654) of 453·79. The self-report model has a null deviance (683) of 505·61 and a residual deviance (682) of 492·74. The amphetamine model explained 4·4% of the variance and the self-report model explained 2·5%, based on McFadden's pseudo-$R^2$.

**Table S5: Mixed-effects logistic models for RQ3 a) without interaction and b) with interaction term**

a)

| Fixed Effects | | | | |
|---|---|---|---|---|
| | Est | SE | z | p |
| (Intercept) | -8·5 | 0·76 | -11·24 | < 0·0001 |
| season | 1·06 | 0·36 | 2·95 | 0·0032 |
| assay | 1·43 | 0·44 | 3·23 | 0·0013 |
| lifetime0 | -0·86 | 1·03 | -0·83 | 0·404 |
| lifetime1 | -0·53 | 0·58 | -0·91 | 0·36 |
| **Random Effects** | | | | |
| | | | Variance | SD |
| individ (Intercept) | | | 60·86 | 7·8 |
| **Model fit** | | | | |
| $R^2$ | | | Marginal | Conditional |
| | | | 0·01 | 0·95 |

b)

| Fixed Effects | | | | |
|---|---|---|---|---|
| | Est | SE | z | p |
| (Intercept) | -9·69 | 1·00 | -9·67 | < 0·0001 |
| season | 1·09 | 0·37 | 2·94 | 0·0033 |
| assay | 2·68 | 0·69 | 3·91 | < 0·0001 |
| lifetime0 | -0·18 | 1·65 | -0·11 | 0·91 |
| lifetime1 | 1·11 | 0·83 | 1·33 | 0·18 |
| assay:lifetime0 | -1·04 | 1·93 | -0·54 | 0·59 |
| assay:lifetime1 | -3·55 | 1·26 | -2·82 | 0·0048 |
| **Random Effects** | | | | |
| | | | Variance | SD |
| individ (Intercept) | | | 72·96 | 8·54 |
| **Model fit** | | | | |
| $R^2$ | | | Marginal | Conditional |
| | | | 0·02 | 0·96 |

Note. N = 682. season = season variable for T1 and T2; assay = immunoassay result; lifetime0 = no lifetime trauma load; lifetime1 = low lifetime trauma load; individ = individual ID; self-report = self-reported khat use last week. Reference categories are negative for the immunoassay test, no use for self-report and high for lifetime trauma load. p-values for fixed effects calculated using Satterthwaites approximations. Confidence Intervals have been calculated using the Wald method. Model equation a): cidi ~ season + assay + lifetime + (1 | individ); b): cidi ~ time + amp300 * trauma_lifetime + (1 | individ)

**Table S6: Type III Wald $\chi^2$ tests for mixed-effects logistic models for RQ3 with interaction term**

| **Effect** | **$\chi^2$** | **df** | ***p*** |
|---|---|---|---|
| (Intercept) | 33·81 | 1 | <0·001 |
| season | 8·64 | 1 | 0·0033 |
| assay | 0·80 | 1 | 0·37 |
| lifetime | 2·04 | 2 | 0·36 |
| assay:lifetime | 7·95 | 2 | 0·019 |

Note. N = 682. season = season variable for T1 and T2; assay = immunoassay result; lifetime = lifetime trauma load.

**Table S7: Pairwise comparisons of immunoassay status within levels of lifetime trauma load (EMMs, log-odds scale) for mixed-effects logistic models for RQ3**

| Trauma level | Contrast | Est | SE | *z*-ratio | *p* |
|---|---|---|---|---|---|
| 0 | assay0 – assay1 | -1·64 | 1·84 | -0·89 | 0·37 |
| 1 | assay0 – assay1 | 0·86 | 0·99 | 0·88 | 0·38 |
| 2 | assay0 – assay1 | -2·68 | 0·69 | -3·91 | 0·0001 |

Note. N = 682. assay0 = negative immunoassay result; assay1 = positive immunoassay result. Trauma levels correspond to ordinal coding of lifetime trauma load variable (0 = none, 1 = low, 2 = high). Results are based on estimated marginal means from a mixed-effects logistic regression model and are reported on the log-odds scale. Analyses are averaged across time.

**Table S8: Binary logistic regression models for dry season T1 for RQ3 with a) no interaction and b) interaction term**

a)

| Predictor | *b* | *SE* | *z* | *p* |
|---|---|---|---|---|
| (Intercept) | -2·38 | 0·24 | --10·07 | < 0·0001 |
| T1_assay | 0·78 | 0·29 | 2·74 | 0·0062 |
| T1_trauma0 | -0·72 | 0·50 | -1·43 | 0·15 |
| T1_trauma1 | -0·47 | 0·30 | -1·56 | 0·12 |

b)

| Predictor | *b* | *SE* | *z* | *p* |
|---|---|---|---|---|
| (Intercept) | -2·47 | 0·27 | -8·89 | < 0·0001 |
| T1_assay | 0·98 | 0·40 | 2·45 | 0·014 |
| T1_trauma0 | -0·51 | 0·65 | -0·77 | 0·44 |
| T1_trauma1 | -0·30 | 0·39 | -0·78 | 0·44 |
| T1_assay:T1_trauma0 | -0·48 | 1·03 | -0·47 | 0·64 |
| T1_assay:T1_trauma1 | -0·38 | 0·61 | -0·63 | 0·53 |

*Note*. N = 668. T1_assay = immunoassay for T1; T1_trauma0 = no trauma load before T1; T1_trauma1 = low trauma load before T1. Reference categories are negative for the immunoassay test, no use for self-report and high for trauma load before T1. *b* represents unstandardized regression weights. For logistic regression these are logits, also named log-odds. The no-interaction model has a null deviance (667) of 384·81 and a residual deviance (664) of 373·46. The interaction model has a null deviance (667) of 384·81 and a residual deviance (662) of 372·96. The no interaction model explained 2·9% of the variance and the interaction model explained 3.1%, based on McFadden's pseudo-$R^2$.

**Table S9: Binary logistic regression models for rainy season T2 for RQ3 with a) no interaction and b) interaction term**

a)

| Predictor | *b* | *SE* | *z* | *p* |
|---|---|---|---|---|
| (Intercept) | -2·37 | 0·22 | -10·61 | < 0·0001 |
| T2_assay | 1·1 | 0·25 | 4·4 | < 0·0001 |
| lifetime0 | -0·93 | 0·55 | -1·71 | 0·088 |
| lifetime1 | -0·23 | 0·26 | -0·9 | 0·37 |

b)

| Predictor | *b* | *SE* | *z* | *p* |
|---|---|---|---|---|
| (Intercept) | -2·59 | 0·28 | -9·02 | < 0·0001 |
| T2_assay | 1·46 | 0·36 | 4·11 | < 0·0001 |
| lifetime0 | -0·54 | 0·78 | -0·7 | 0·49 |
| lifetime1 | 0·21 | 0·39 | 0·53 | 0·597 |
| T2_assay: lifetime0 | -0·68 | 1·09 | -0·62 | 0·54 |
| T2_assay: lifetime1 | -0·78 | 0·53 | -1·47 | 0·14 |

*Note.* N = 656. T2_assay = immunoassay for T2; lifetime0 = no lifetime trauma load; lifetime1 = low lifetime trauma load. Reference categories are negative for the immunoassay test, no use for self-report and high for lifetime trauma load. *b* represents unstandardized regression weights. For logistic regression these are logits, also named log-odds. The no-interaction model has a null deviance (655) of 474·51 and a residual deviance (652) of 450·02. The interaction model has a null deviance (655) of 474·51 and a residual deviance (650) of 447·74. The no interaction model explained 5·2% of the variance and the interaction model explained 5.6%, based on McFadden's pseudo-$R^2$.

**Table S10: Binary logistic regression model for rainy season T2 for RQ3 with three-way interaction among participants who experienced trauma load before T1**

| Predictor | *b* | *SE* | *z* | *p* |
|---|---|---|---|---|
| (Intercept) | -2·4 | 0·30 | -7·95 | < 0·0001 |
| T2_assay | 0·41 | 0·46 | 0·87 | 0·38 |
| T1_trauma2 | -0·43 | 0·51 | -0·82 | 0·41 |
| T2_recent_trauma | -0·33 | 0·66 | -0·5 | 0·62 |
| T2_assay:T1_trauma2 | 1·13 | 0·69 | 1·62 | 0·11 |
| T2_assay: T2_recent_trauma | 1·41 | 0·83 | 1·7 | 0·0898 |
| T1_trauma2: T2_recent_trauma | 1·12 | 0·90 | 1·24 | 0·21 |
| T1_trauma2: T2_recent_trauma: T2_assay | -2·0 | 1·13 | -1·76 | 0·079 |

Note. N = 570. T2_assay = immunoassay for T2; T1_trauma2 = high trauma load before T1. Reference categories are negative for the immunoassay test, no use for self-report and low for trauma load before T1 as well as no recent trauma. *b* represents unstandardized regression weights. For logistic regression these are logits, also named log-odds. The model has a null deviance (569) of 428·54 and a residual deviance (562) of 402·12. The model explained 6·2% of the variance, based on McFadden's pseudo-$R^2$.
